# Clearing the Fog: A Systematic Review on Cognitive Dysfunction in COVID-19

**DOI:** 10.1101/2022.05.24.22275552

**Authors:** Nicole D. Butardo, Mikaela Frances D. Coronel, Alanna Marie O. Dino, Tiffany Ritz F. Mendoza, Oliver Kyle DC. Sto. Domingo, Zypher Jude G. Regencia, Jacqueline C. Dominguez, Emmanuel S. Baja, Antonio D. Ligsay

**Affiliations:** College of Science, University of Santo Tomas, España Blvd., Manila 1008 Philippines; Department of Clinical Epidemiology, College of Medicine, University of the Philippines-Manila, Pedro Gil Street, Ermita, Manila, 1000 Philippines; Institute of Clinical Epidemiology, National Institutes of Health, University of the Philippines-Manila, Pedro Gil Street, Ermita, Manila, 1000 Philippines; Institute for Neurosciences, St. Luke’s Medical Center, 279 E. Rodriguez Sr. Ave, Quezon City, 1112 Philippines; The Graduate School, University of Santo Tomas, España Blvd., Manila 1008 Philippines; St. Luke’s Medical Center College of Medicine - William H. Quasha Memorial, 279 E Rodriguez Sr. Ave, Quezon City, 1112 Metro Manila

**Keywords:** brain fog, cognitive dysfunction, COVID-19, disease severity, neuroinflammatory processes, pathophysiology, SARS-CoV-2 infection

## Abstract

**Objective:** The systematic review aims to examine the association between COVID-19 and cognitive dysfunction, including the link between the severity of COVID-19 and the occurrence of cognitive impairment and the potential pathophysiological mechanisms related to brain fog among COVID-19 patients.

**Methods:** PubMed, Oxford University Press, ProQuest Health and Medical Complete, ScienceDirect, Ovid, HERDIN, Google Scholar, and Cochrane Library databases were accessed to retrieve literature using the PRISMA guidelines.

**Results:** After critical appraisal, thirteen full journal articles were included in the study. The studies showed the most frequent cognitive impairment are attention, memory, and executive function in COVID-19 patients. Compared with healthy controls (HC) in 3 out of 4 studies, cognitive impairment was only evident in COVID-19 patients. Furthermore, two studies showed no correlation between brain fog and depression, and five studies showed a link between the severity of COVID-19 infection and cognitive impairment. Cases ranging from mild to severe illness presented manifestations of brain fog. However, a disparity in the evidence of the pathophysiology of COVID-19 and cognitive dysfunction exists, prompting the need to investigate further. Additionally, recent studies provide insufficient evidence for direct central nervous system invasion, and there are emerging studies that contrast the presumed pathogenesis of neurological complications from neuroinflammation.

**Conclusion:** There is an association between COVID-19 and cognitive dysfunction. Manifestation of cognitive dysfunction is present regardless of illness severity. Moreover, there are existing pathophysiological mechanisms of the Coronavirus that lead to cognitive dysfunction in COVID-19 patients; however, additional studies are required to substantiate such mechanisms further.

**PROSPERO Registration Number:** CRD42022325669

## INTRODUCTION

With millions of Coronavirus (COVID-19) cases worldwide, it is becoming apparent that more people are experiencing neurological symptoms associated with severe acute respiratory syndrome coronavirus-2 (SARS-CoV-2) infection. For this reason, researches into the putative link between SARS-CoV-2 and neurological manifestations continue to grow (Desai et al., 2021; Ghannam et al., 2020; Guadarrama-Ortiz et al., 2020). Cognitive dysfunction, also known as brain fog, is defined as the decrements in the cognitive status during continuous mental activity (García-Sánchez et al., 2022). Brain fog as a general term can present as confusion, difficulty finding the appropriate words, disorientation, memory problems, altered mental status, and trouble concentrating (Altuna et al., 2021; Asadi-Pooya et al., 2022; Hampshire et al., 2021). Such decline can be measured using various neurocognitive assessments that can either reveal a cognitive impairment with a certain severity or discern which specific cognitive domain is significantly affected. According to the Montreal Cognitive Assessment (MoCA) test, the cognitive domains include orientation, attention, language, visuospatial function, memory, and executive function (Julayanont & Nasreddine, 2017).

Gustatory and olfactory impairments are the most frequent sudden neurologic manifestations of COVID-19 associated with the peripheral nervous system (PNS), which occur in the early stages of SARS-CoV-2 infection (Cooper et al., 2020; Lechien et al., 2020; Salcan et al., 2021). These sensory impairments entail that SARS-CoV-2 is afflicting the nervous system (Iadecola et al., 2020). The SARS-CoV-2 virus could potentially affect the gustatory system in two different ways– either by directly damaging the mucosal membrane of the oral cavity and the peripheral neuronal trajectory of the gustatory tract or by directly damaging the cranial nerves that are responsible for the sense of taste (Finsterer & Stollberger, 2020). With the presentation of anosmia in COVID-19 patients, olfactory neurons were at an incredibly high risk of injury due to the high viral load within the nasal cavity (Zou et al., 2020).

The discovery of the brain invasion of mice by the intranasal administration of the novel coronavirus (Kumari et al., 2021) and the existing knowledge about the previous detection of other coronaviruses in human cerebrum (Cheng et al., 2020) led to a hypothesis that COVID-19 infection can be linked with neurocognitive complications. Preliminary studies suggest that cognitive deficits of hospitalized COVID-19 patients were dependent on two factors: the medical assistance they received (Ferrucci et al., 2021; Hampshire et al., 2021) and the degree of inflammation (Zhou et al., 2020); that is, severe infections are assumed to contribute to severe cognitive impairments (Beaud et al., 2021; Tay et al., 2021; Wild et al., 2021). However, there are still gaps in the studies regarding how COVID-19 infection increases cognitive impairment risk, severity, and progression. There is mounting evidence that brain fog is among the observed neurological manifestations of COVID-19 (Whittaker et al., 2020). However, the association of the symptom, including the possible mechanisms and the potential triggers, remains unclear. Consequently, if there is an association between the two, research about the extent of the impact on cognition and the affected cognitive domains of brain fog is limited (Zhou et al., 2020). Therefore, researchers have yet to determine whether cognitive systems are equally affected or some domains are more susceptible to SARS-CoV-2 infection. The primary purpose of this review is to assess cognitive dysfunction, more commonly known as “brain fog,” as a symptom of COVID-19 to understand its etiology better. This encompasses the mechanisms that may cause the impairment and the prevalence of the emergent symptom. The current study also aims to determine the association between SARS-CoV-2 infection and cognitive dysfunction, elucidate the link between the severity of COVID-19 infection and brain fog, and describe the potential pathophysiological mechanisms related to cognitive dysfunction in COVID-positive individuals. This study gathered evidence that could provide clarity on the association between brain fog and COVID-19 infection.

## METHODS

### Study Setting

The study was carried out for six months (July to December 2021). Database searching was the initial step for identifying reports. Search strategies were employed, and primary identified records were based on the titles, database availability, and abstracts. Reviewers evaluated these based on predetermined inclusion criteria. Once reports have passed the inclusion criteria, a full-text report was obtained to assess its eligibility against the inclusion criteria further. Irretrievable full-text reports were excluded. More specifically, a population, phenomenon of interest, and context (PICO) approach was utilized to generate a sequence of terms (Lockwood et al., 2015; Munn et al., 2018).

### Eligibility Criteria/ Criteria for Considering Studies for this Review

#### Types of Studies

The studies included in this systematic review were observational studies, including case reports, case series, cross-sectional studies, case-control studies, and cohort studies. In addition, reports focusing on individuals with COVID-19 who suffer from brain fog were included. Only articles in the English language restriction were imposed.

#### Types of Participants

Participants included in the study were patients who tested positive for COVID-19 through a Reverse Transcription Polymerase Chain Reaction (RT-PCR), antigen test or SARS-CoV-2-specific immunoglobulin G (IgG) in serum and experienced cognitive dysfunction. All COVID-19 positive patients were included regardless of sex, race, ethnicity, and age. In addition, healthy, non-COVID-19 patients were also included for comparison.

#### Types of Phenomena of Interest and Context

This systematic review’s main phenomena of interest included the manifestation of brain fog in COVID-19 patients and non-COVID-19 patients. Therefore, studies focusing on the link between COVID-19 severity and occurrence of brain fog and potential mechanisms that cause cognitive dysfunction as a symptom of COVID-19 were also included.

### Search Methods for Identification of Studies

#### Information Sources

The following databases were used to identify completed studies until December 2021: PubMed, Oxford University Press, ProQuest Health and Medical Complete, ScienceDirect, Ovid, HERDIN, Google Scholar, and Cochrane Library. Ongoing studies were recognized; however, these studies were not included in the systematic review. In addition, a date restriction was set for identifying studies: studies since the start of the COVID-19 pandemic, January 2020 up to December 2021, were included in this systematic review. Furthermore, English language restriction was imposed.

#### Search Strategy

The electronic literature search included the following key terms: COVID-19, SARS-CoV-2, neurocognitive impairment, brain fog, confusion, poor concentration, memory problems, brain fog pathophysiology, neurological mechanism, and cognitive dysfunction. Moreover, the Boolean search strategy (“AND,” “NOT,” “AND NOT,” “OR”) was employed to identify studies using the key terms. The search strategy for databases can be found in online Supplemental Appendix A.

#### Selection Process

Authors independently searched for studies that were included in the systematic review. The initial selection of studies consists of the examination of the titles, abstracts, and full-text, if available. Inclusion criteria implemented were observational studies on individuals with COVID-19 who experience cognitive dysfunction. The authors created the final list of included studies. Any disagreements that arose during the appraisal process were settled by discussing with another author. *Assessment of Risk of Bias in Individual Studies*

Three authors (MFDC, TRFM, and OKSD) independently evaluated the risk of bias in each study. A fourth (NDB) and fifth (AMOD) author was assigned to resolve disagreements in assessments. To assess bias in included studies, they were segregated into observational studies such as case reports, case series, cross-sectional studies, case-control studies, and cohort studies. The risk of bias in the included studies was assessed using the Strengthening the Reporting of Observational Studies in Epidemiology (STROBE) statement (Vandenbroucke et al., 2007). The risk of bias was scored as “low,” “moderate,” or “high” risk. The overall quality of each study was given a rating based on the score from the STROBE statement. Furthermore, only studies with a low-risk rating were included in the study.

## Data analysis and presentation

Formatted according to the extraction tool used to extract the data, an MS Excel spreadsheet was used to tabulate results for representation of results. The characteristics of the included articles were described, which were previously discussed and agreed upon within the study team.

## Evidence Synthesis

A narrative synthesis of overall evidence was undertaken by comparing and contrasting the data to express and synthesize the results of the included studies. Development of a preliminary synthesis, exploration of the relationships within and between studies, and the determination of the robustness of the synthesis were the three stages of the narrative synthesis undertaken by the research team (Popay et al., 2006). Data of the included studies were qualitatively described and presented. The authors frequently met to discuss the results and reach a consensus on the findings.

## RESULTS

### Description of the Studies

#### Literature Search

Two hundred eighty-nine studies were identified after a comprehensive search through databases (PubMed, ProQuest, Oxford University Press, ScienceDirect, Cochrane Library, Google Scholar, JSTOR, and Herdin). Duplicate records (n=12) were removed based on their titles. After initial screening, 177 studies were excluded due to differences in study design (*e*.*g*., review paper, narrative review, hypothesis/theory article and chart review) based on the title and abstract. In addition, 15 articles were not retrieved because the study is ongoing. A total of 85 full-text studies were assessed for eligibility. After critical appraisal, 13 studies were included in the final systematic review. An adapted PRISMA (Preferred Reporting Items for Systematic Reviews and Meta-Analyses) flow chart of the study selection is presented in Fig. 1.

**Figure 1.**
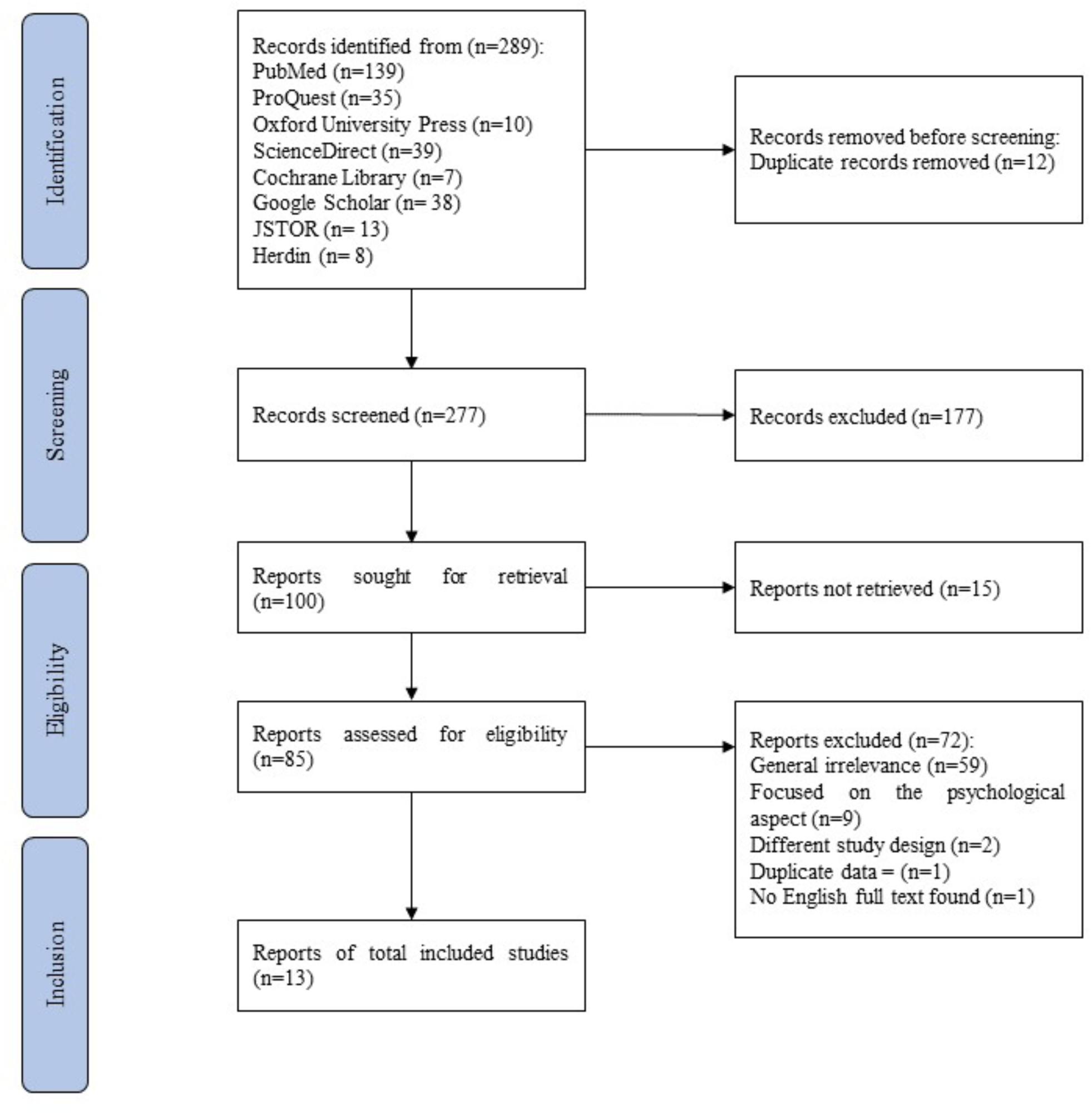
PRISMA flow diagram.

#### Included Studies

Characteristics of the included studies are tabulated in Table 1. The age of the study participants in the included studies was greater than 16 years old. All study participants were diagnosed with COVID-19 through various diagnostic tests such as SARS-CoV-2 RT-PCR of a nasopharyngeal swab, SARS-CoV-2 antibody testing (e.g., SARS-CoV-2-specific IgG in serum), rapid antigen test, and polymerase chain reaction (PCR) test for SARS-CoV-2 in upper and/or lower airway samples. Participants in 4 included studies were assessed during COVID-19 infection, seven during post-COVID-19 infection, 4 had follow-ups, and one examined patients post-mortem. The duration of the follow-ups in the included studies ranges from 2 to 4 months. Out of the 13 included observational studies, 1 is a case series, 1 is a case report, 6 were cross-sectional studies, and 5 were cohort studies.

**Table 1.** Characteristics of included studies (n=13).

| Author<br>(Year) | Sample<br>Size | Population<br>Mean Age/<br>Median Age*<br>(Range) | Study<br>Design | Outcome/s |
| --- | --- | --- | --- | --- |
| Beach et al.<br>(2020) | 4 | 75.25 yo (68-87) | Case series | Neuropsychological and neurophysiological features of fatigue were studied in post-COVID-19 patients. In addition, apathy, deficits in executive functions, and reduction in global cognition were found. |
| Blazhenets et al. (2021) | 31 | 66.00 yo (39-89) | Cohort | MOCA $M = 19.13$; 23.38<br>Impaired language, visuospatial function, memory, and executive function |
| Garcia, M.A. et al. (2021) | 18 | 56 yo* (20-79 yo; 14 healthy, 68 non-COVID-19 neurological disease controls) | Cross-sectional | Neuroinflammatory processes in COVID-19 CSF may be the result of a homeostatic neurological response rather than an adaptive, immune-mediated cytokine storm or inflammation caused by neurovirulence.<br>Mild, moderate, severe, and critical cases exhibited cognitive deficits, regardless of severity. |
| Garcia, M.H.C. et al. (2021) | 199 | 43 yo (>18 yo) | Cross-sectional | Mild to moderate cases of COVID-19 severity exhibited cognitive deficits |
| Graham et al. (2021) | 100 | 43.2 yo | Cohort | Patients and controls had mild to moderate cognitive dysfunction but showed no significant difference |
| Hosp et al. (2021) | 29 | 65.2 yo (>18) | Cohort | MOCA $M = 21.77$<br>Impaired attention, visuospatial function, memory, and executive function |
| Ortelli et al. (2020) | 12 | 67 yo (48-80) | Cross-sectional | MOCA $M = 17.80$<br>Impaired attention via SIT and NT but unimpaired attention on VT |

| **Author**<br>**(Year)** | **Sample**<br>**Size** | **Population**<br>**Mean Age/**<br>**Median Age\***<br>**(Range)** | **Study**<br>**Design** | **Outcome/s** |
| --- | --- | --- | --- | --- |
| Puchner et al. (2021) | 23 | 57 yo ($\geq 18$) | Cohort | In 29% of tested patients, cognitive deficits in concentration, memory, and/or executive functions were found. |
| Vannorsdall et al. (2022) | 82 | 54.5 yo ($>16$) | Cohort | Impaired scores for Oral Trail Making Test, Verbal Fluency, and Rey Auditory Verbal Learning Test<br>Critically ill patients exhibited cognitive deficits. |
| Virhammar et al. (2021) | 19 | 64 yo (34-76) | Cohort | Majority of patients presented with increased CSF levels of neuronal injury markers.<br>Results of the study show that there is no specific pattern in the manifestation of brain fog across illness severity. |
| Woo et al. (2020) | 18 | 42.11 yo (17-71) | Cross-sectional | TICS-M: COVID-19 patients scored significantly lower than HC, especially in memory, attention, and language. Patients with mild to moderate case severity indicated manifestation of cognitive dysfunction. |
| Yesilkaya et al. (2021) | 1 | 20 yo | Case report | Cognitive deficits in SARS-CoV-2 infection can result from glutamatergic dysfunction with decreased glutamate and NAA levels in the DLPFC confirmed by MRS. |
| Zhou et al. (2020) | 29 | 47 yo (30-64) | Cross-sectional | CPT: COVID-19 patients performed significantly worse than HC<br>No significance for TMT, SCT, and DST between the two groups |

#### Excluded Studies

A total of 276 studies were excluded from the list of included studies due to the following reasons: different study design (n=179), irrelevance to the systematic review (n=59), unavailability of the full-text article (n=16), duplications (n=13), and focused on the psychological aspect (n=9).

### Risk of Bias in Included Studies

Observational studies were evaluated based on ten domains: Introduction - (1) Objectives; Methods - (2) Participants; Results - (3) Participants, (4) Descriptive Data,(5) Outcome Data, and (6) Main Results; Discussion - (7) Key Results, (8) Limitations,(9) Interpretation, and (10) Generalizability. Of the 13 included observational studies, all were assessed as low risk of bias.

### Qualitative Results

Eleven studies focused on the association between COVID-19 and brain fog, five (5) studies identified the link between the severity of COVID-19 and the occurrence of cognitive dysfunction, and seven (7) studies described the possible pathophysiological mechanisms related to brain fog in COVID-19 patients.

*Association between SARS-CoV-2 infection and neurocognitive dysfunction* The prospective cohort study conducted two MoCA tests with mean scores of 19.1 and 23.4 on the first and second exams, respectively (Blazhenets et al., 2021).

The average scores showed significant improvement on the second exam. However, the MoCA performance of the second exam was still within the range of mild cognitive impairment (MCI). In a cross-sectional study, the average MoCA score of the COVID-19 patients was significantly lower than the healthy controls (Ortelli et al., 2021). In addition, the severity of cognitive impairment was mentioned in a study(Hosp et al., 2021) wherein 54% (14/26) of the participants were mild to moderately impaired, whereas 15% (4/26) were severely impaired (see Table 2 for details).

**Table 2.** Summary of domain-specific and non-domain-specific test results of included studies (n=7).

| Author | Sample Size | MoCA global score (> 26) |  | Cognitive Domains* |  |  |  |  |  |
| --- | --- | --- | --- | --- | --- | --- | --- | --- | --- |
|  |  |  |  | O | A | L | VSF | M | EF |
| Blazhenets et al. (2021) | 8 | 1st exam | 2nd exam | x | x | ✓ | ✓ | ✓ | ✓ |
|  |  | 19.13 | 23.38 |  |  |  |  |  |  |
| Graham et al. (2021) | 50 | n/a |  | n/a | ✓ | n/a | n/a | ✓ | ✓ |
| Hosp et al. (2021) | 26 | 21.77 |  | x | ✓ | x | ✓ | ✓ | ✓ |
| Ortelli et al. (2020) | 12 | 17.80 |  | n/a | ✓ | n/a | n/a | n/a | ✓ |
| Vannorsdall et al. (2021) | 82 | n/a |  | n/a | x | ✓ | n/a | ✓ | ✓ |
| Woo et al. (2020) | 18 | n/a |  | ✓ | ✓ | ✓ | n/a | ✓ | n/a |
| Zhou et al. (2020) | 29 | n/a |  | n/a | ✓ | n/a | x | x | x |

\*✓: impaired domain; X: unimpaired domain; n/a: no available data.
**Legend:** O – Orientation; A – Attention; L – Language, VSF – Visuospatial Function; M – Memory; EF – Executive Function

Seven of the included studies reported domain-specific cognitive deficits. The prospective cohort assessed all six (6) cognitive domains that showed an insignificant decline in orientation and attention domains with 6.00 and 5.13 scores, respectively (Blazhenets et al., 2021). However, the MoCA domain scores on language (3.88), visuospatial function (3.13), memory (2.25), and executive function (2.50) revealed significant cognitive deficits.

Another cohort study also utilized the MoCA test (Hosp et al., 2021). The MoCA domain scores in this study revealed an impairment in executive function, visuospatial function, memory, and attention. The two other domains, language, and orientation, were not impaired. An extended neuropsychological test battery confirmed the deficit in executive function and memory, but not in attention (Hosp et al., 2021).

In a cross-sectional study, two cognitive domains such as attention and executive function, were assessed using three subtests of MoCA and Frontal Assessment Battery (FAB), respectively (Ortelli et al., 2021). The reaction time (RT) of the COVID-19 patients in each subtest of MoCA was compared with that of the healthy controls. In two of the subtests, namely Stroop Interference Task (SIT) and Navon Task (NT), the RTs were significantly longer in the patients than in the healthy controls (HC). In contrast, the RTs of the patients and the healthy controls in the Vigilance Task (VT) did not have a significant difference. Furthermore, significantly lower FAB scores were observed in COVID-19 patients than in the healthy controls indicating an executive function deficit (Ortelli et al., 2021).

Another study also utilized subtests of MoCA such as the Trail Making Test (TMT), Sign Coding Test (SCT), Continuous Performance Test (CPT), and Digit Span Test (DST) that revealed significant differences in the attention domain and no significant differences in executive function, visuospatial function, and memory between COVID-19 patients and healthy controls (Zhou et al., 2020). Meanwhile, in a cohort study, a neuropsychological assessment battery revealed specific cognitive deficits in non-ICU and post-ICU COVID-19 patients (Vannorsdall et al., 2022). Both groups showed significant cognitive impairment in three cognitive domains, namely executive function, language, and memory but not in attention.

A different cognitive function assessment tool was used in another cohort study (Graham et al., 2021). In this study, the National Institutes of Health (NIH) Toolbox v2.1 instrument revealed a mild to moderate cognitive impairment in both COVID-19 patients and healthy controls; however, it did not show any significant difference between the Toolbox T-scores of the two groups.

A Modified Telephone Interview for Cognitive Status (TICS-M) is another screening tool for MCI used in another study (Woo et al., 2020). Cognitive deficits in orientation, memory, attention, and language in post-COVID-19 patients and healthy controls were reported. It was also revealed that the scores of the patients in three domains, specifically memory, attention, and language, were significantly lower than the healthy controls (Woo et al., 2020).

Moreover, a cross-sectional study involving 199 patients from Lima, Peru, described the relationship between mild to moderate COVID-19 infection and neurological symptoms, wherein 7 out of 199 patients (3.5%) presented impaired consciousness (M. H. C. Garcia et al., 2021). Meanwhile, in another study, 23 patients who had severe to critical COVID-19 were analyzed (Puchner et al., 2021). A neuropsychological evaluation was conducted on 14 out of the 23 patients, and four patients (29%) were found to have cognitive dysfunction in memory, executive function, and attention (Puchner et al., 2021). In addition, a case study reported a patient who tested positive for SARS-CoV-2 and did not have any neurologic or psychiatric evaluation history. The patient was evaluated using a neuropsychological battery test, including the Global Deterioration Scale (GDS), California Verbal Learning Test (CVLT), FAB, and TMT. The neurological battery tests suggested cognitive impairment (FAB score of 16; GDS stage 3) in executive function, concentration, and memory. At the patient’s follow-up, no cognitive impairment was identified (FAB score of 13; GDS stage 1) (Yesilkaya et al., 2021).

Fourteen COVID-19 patients underwent hospital anxiety and depression scale (HADS-D) that revealed no significant increase in their anxiety and depression symptoms despite detecting cognitive deficits in memory and executive functions.(Puchner et al., 2021) A similar study that utilized the Patient Health Questionnaire - 9 (PHQ-9) Depression Scale revealed no significant correlation between depression and cognitive dysfunction.(Woo et al., 2020) Meanwhile, one study used the Beck Depression Inventory (BDI), wherein scores of COVID-19 patients and healthy controls had a significant difference (Ortelli et al., 2021).

#### Elucidating the link between disease severity of COVID-19 infection and the manifestation of cognitive dysfunction

The National Institute of Health (NIH) established a classification for the clinical spectrum of SARS-CoV-2 infection. Patients with COVID-19 may be grouped according to illness severity: asymptomatic or presymptomatic, mild, moderate, severe, and critical illness (Health, 2020). Increasing evidence suggests that the severity of SARS-CoV-2 infection is linked with the occurrence of cognitive dysfunction in COVID-19 patients. After evaluation of retrieved and appraised journals, five studies correlate with this hypothesis (see Table 3). In the cross-sectional study examining cerebrospinal fluid (CSF) of 18 COVID-19 subjects with neurological complications, 44% (8) were classified as critical, 28% (5) as severe, 22% (4) as moderate, and one patient as mild illness (M. A. Garcia et al., 2021). This review shows that across the case of 18 COVID-19 patients, all exhibited cognitive dysfunction regardless of case severity. Furthermore, a cohort study conducted among patients of the John Hopkins Post-Acute COVID-19 Team (PACT) Pulmonary Clinic reveals that out of 82 patients classified for critical illness, 67% (54) demonstrated abnormally low cognitive scores (≥1 deviation from published age-adjustive normative means) and is correlated to mild/moderate or severe range of cognitive impairment (Vannorsdall et al., 2022). In addition, the prospective-single center study involved 19 patients with a distribution of 11% (2) having a mild illness, 21% (4) having a moderate illness, another 21% (4) having a severe illness, and 47% (9) having critical illness severity (2021) were reported (Virhammar et al., 2021). Report also showed that 2 out of 9 critically ill patients manifested cognitive dysfunction, 2 out of 4 severely ill patients showed altered mental status, and 2 out of 4 moderately ill patients exhibited confusion and altered mental status (Virhammar et al., 2021). Findings show that there is no specific pattern for COVID-19-related cognitive dysfunction. Lastly, a cross-sectional study conducted at the University Medical Center Hamburg-Eppendorf involved 18 patients in either mild or moderate severity (Woo et al., 2020). Cognitive deficiencies were present among the involved patients: 9 (50%) reported having attention deficiency, 8 (44.4%) suffered from concentration deficits, 8 (44.4%) experienced short-term memory deficiency, and 5 (27.8%) had trouble finding words. Results from 18 patients impacted with COVID-19 were compared to 10 healthy non-COVID patients. Findings also show that COVID-19 neurological sequelae are independent of hospitalization and illness severity (Woo et al., 2020).

**Table 3.** Summary of occurrence of cognitive dysfunction across illness severity from included studies (n=5).

| Author<br>(Year) | Clinical Spectrum of SARS-CoV-2 Infection |  |  |  |  | Cognitive<br>dysfunction |
| --- | --- | --- | --- | --- | --- | --- |
|  | Asymptomatic or<br>Presymptomatic<br>Infection | Mild<br>Illness | Moderate<br>Illness | Severe<br>Illness | Critical<br>Illness |  |
| Garcia MA et al. (2021) | X | ✓ | ✓ | ✓ | ✓ | ✓ |
| Garcia MHC et al. (2021) | X | ✓ | ✓ | X | X | ✓ |
| Vannorsdall et al. (2022) | X | X | X | X | ✓ | ✓ |
| Virhammar et al. (2021) | X | ✓ | ✓ | ✓ | ✓ | ✓ |
| Woo et al. (2020) | X | ✓ | ✓ | X | X | ✓ |

#### Possible pathophysiological mechanisms leading to cognitive dysfunction

Seven studies analyzed cognitive dysfunction in COVID-19 patients on a molecular level and their potential pathogenesis. One study examined serum pro-inflammatory markers (interleukin-2 (IL-2), IL-4, IL-6, IL-10, tumor necrosis factor-α (TNF-α), interferon-γ (IFN-γ), and C-reactive protein (CRP)) (Zhou et al., 2020). However, their findings revealed no significant correlation between the inflammatory markers and the cognitive function assessment results from the Trail Making Test, Sign Coding Test, and Digital Span Test. In contrast, all the 29 post-COVID-19 patients showed a trend of significant difference for lower reaction time in the first and second parts of CPT and a lower correct number in the second part of CPT than the 29 healthy controls. The serum C-reactive protein level and reaction time in the Continuous Performance Test are positively correlated. Moreover, 11 post-COVID-19 patients demonstrated hyper-inflammation, evidenced by elevated C-reactive protein (CRP) and interleukin-6 serum levels (Ortelli et al., 2021). The remarkable pro-inflammatory state induced by SARS-CoV-2 in all 4 cases and the cessation of carbidopa-levodopa in one patient case described in another study has been postulated to convey a dysregulated immune response and a potential precipitant of the abrupt mental confusion and emotional disarray (Beach et al., 2020).

One study analyzed the CSF of 19 patients with mild to critical COVID-19. Only one (5%) was positive for SARS-CoV-2. The analysis includes biomarkers of central nervous system injury (neurofilament light chain (NfL) protein, glial fibrillary acidic protein (GFAp), and total tau (T-tau), and found increased CSF levels of NfL (63%), total tau (37%), and GFAp (16%). The amount of CSF NfL was higher in patients with central neurological symptoms, and the elevated level was associated with the severity of the disease, time spent in critical care, and level of consciousness (Virhammar et al., 2021).

Another study evaluated fluorodeoxyglucose (FDG)-positron emission tomography (PET) images of 8 COVID-19 patients (post-infection) and the MoCA scores to assess neuronal damage or synaptic dysfunction distribution (Blazhenets et al., 2021). They discovered that after the patients are no longer infectious and in the chronic stage (about six months after symptom onset), their impaired neocortical glucose metabolism can return to normal levels, evidence of reversibility, which is critical for the pathophysiology of cognitive deficit. In the FDG PET scans of another study, 10 of the 15 patients had abnormal findings (Hosp et al., 2021). According to the observer, 10 of the subjects had cortical hypometabolism, with 2 cases having striatal hypermetabolism. Likewise, the second observer noted cerebral hypometabolism in 6 out of 9 subjects. Striatal hypermetabolism is present in the other three cases. In addition, their findings demonstrated a highly significant linear association between MoCA and PET, with more robust pattern expression being related to poorer cognitive function. The same study found oligoclonal bands present with identical electrophoretic patterns in the serum in one patient but unremarkable protein and IgG levels in 4 out of 29 who agreed to undergo CSF analysis. Moreover, all CSF samples tested negative for SARS-CoV-2 (Hosp et al., 2021).

In a case study, the CSF analysis through lumbar puncture is unremarkable. The N-acetylaspartate (NAA), glutamate, and glutamate/glutamine ratio were measured using MR-spectroscopy and bilateral DLPFC. A week and three months following the initial diagnosis of SARS-CoV-2 infection, substantial increases in glutamate, glutamine, and NAA levels were discovered during follow-up, suggesting that the glutamatergic pathway might be implicated in the pathogenesis of cognitive impairment (Yesilkaya et al., 2021).

## DISCUSSION

All studies that utilized the MoCA test showed MCI following COVID-19 infection. An included study reported a more severe global cognitive impairment among COVID-19 patients than healthy controls (Ortelli et al., 2021). Among the cognitive domains, attention, executive function, and memory are most likely to be impaired. These were also the domains that frequently showed significant differences between COVID-19 and non-COVID-19 patients in a recent study (Crivelli et al., 2022). This observation is congruent with another study reporting that the exact cognitive domains, except for memory, seem prone to impairments (Daroische et al., 2021). On the other hand, results on language, orientation, and visuospatial function varied in the included studies. This inconsistency may be due to the apparent heterogeneity in the tests used, time of evaluation, eligibility criteria, and the presentation of results.

Majority of the included studies utilized the MoCA test (n = 5), followed by NIH Toolbox (n = 1) and TICS-M (n = 1). Different tests with varying total scores, sensitivity, and specificity in comparing study populations can be attributed to the contradictory results. For instance, one study used three tests, SIT, NT, and VT, all assessed the attention domain and discovered that only SIT and NT showed significantly different RTs between COVID-19 and healthy controls (Ortelli et al., 2021). However, the difference in VT was not statistically different. Furthermore, some studies either did not specify or utilized different cut-off scores. Two studies used the same test and total score. Still, different cut-off values in assessing visuospatial function, thereby affecting the consistency of the interpretation (Blazhenets et al., 2021; Hosp et al., 2021). This emphasizes the need for a standardized and accurate test specific to each cognitive domain.

A prospective cohort study with 31 participants showed a better MoCA performance of COVID-19 patients at the chronic stage than at the subacute stage (Blazhenets et al., 2021). The authors provided evidence that supports the association between COVID-19 infection and cognitive dysfunction, and these deficits can still be measured six months after symptom onset of COVID-19. However, only eight patients underwent a second MoCA examination at the chronic stage of the disease. Similarly, another study reported that only one-tenth of the patients had cognitive dysfunction at the 6-month follow-up (Nalbandian et al., 2021). With that, the different assessment times of the included studies could be the result of the inconsistency. More research using the same methodology is necessary to evaluate the time frame of cognitive dysfunction after COVID-19 infection.

Several studies showed the comparison of results between COVID-19 and non-COVID-19 patients (healthy controls). Three of the four studies used HC and reported a cognitive impairment only in COVID-19 patients (Ortelli et al., 2021; Woo et al., 2020; Zhou et al., 2020). A similar review reported that its included studies had significantly more cases of cognitive impairment in COVID-19 patients than in HC (Daroische et al., 2021).

Of all included studies, only three studies measured depression, two of which stated no correlation between depression and cognitive dysfunction. However, one study reported a significant difference in depression between COVID-19 patients and HC (Ortelli et al., 2021). Previous studies hypothesized that cognitive dysfunction and depression was bi-directional (Miskowiak et al., 2021; Vinkers et al., 2020). Severe cognitive dysfunctions may produce more depression because of increased difficulty in daily life functions. In addition, more symptoms of depression can affect the performance in cognitive tests (Miskowiak et al., 2021). Therefore, there should be more significant consideration of mood and cognitive symptoms following COVID-19 infection.

Emerging reports indicate that a large population suffers from cognitive dysfunction due to COVID-19 infection. Results from the selected studies (Table 3) showed that cognitive dysfunction is present among patients characterized by mild, moderate, severe, and critical illnesses. According to a report, cognitive impairments were most prevalent in hospitalized patients (Hampshire et al., 2021). However, non-hospitalized patients also exhibited cognitive dysfunction relating to COVID-19 infection. A recent study stated evidence of cognitive deficits in patients classified with mild to moderate illness (Del Brutto et al., 2021). Cases exhibiting severe illness also provide proof of cognitive deficit (Hampshire et al., 2021; Negrini et al., 2021).

Additionally, severe infections are assumed to contribute to severe cognitive impairments (Beaud et al., 2021; Tay et al., 2021; Wild et al., 2021). Moreover, another study reported that critically ill patients presented long-lasting complaints of inability to think and concentrate (brain fog) (Asadi-Pooya et al., 2022). In contrast with the current study results, other studies revealed that cognitive deficits were also observed in asymptomatic/presymptomatic cases (Amalakanti et al., 2021; Huang et al., 2021; Tenforde et al., 2020). Therefore, regardless of the intensity of clinical manifestations of COVID-19, patients may still develop brain fog; however, take into account that the link between the severity of COVID-19 and severity of brain fog was not elucidated. The correlation between the severity of COVID-19 disease & severity of cognitive dysfunction remains inconclusive due to a lack of studies, variation of patient characteristics, and breadth and depth of cognitive assessment. The establishment of standardized tests for obtaining cognitive scores should be developed to represent a normative data set to analyze the correlation of severity scale.

Out of 13 studies, only three tested for the presence of SARS-CoV-2 in the CSF. No evidence of viral RNA in the CSF was reported (M. A. Garcia et al., 2021). Likewise, all tested negative among 4 out of 29 patients who underwent lumbar puncture (Hosp et al., 2021). On the other hand, 1 out of 19 patients tested positive for SARS-CoV-2 in CSF findings (Virhammar et al., 2021). The lack of studies reporting the presence of viral RNA in the CSF leads to inadequate evidence of SARS-CoV-2 neuroinvasion, suggesting that it is not the primary pathogenic mechanism in most cases. The NfL protein is an indicator of neuroaxonal damage. Recent research of 544 Mexican Americans found that NfL had a detrimental influence on processing speed, attention, executive skills, and delayed recognition memory in normal and mild cognitive impairment groups, suggesting its significance as a marker of cognitive impairment and early cognitive impairment changes (Hall et al., 2020). In a previous study that used a community-based population of non-cognitive impairment participants, CSF NfL is a better predictor of cognitive deterioration than other CSF markers of neurodegeneration (Mielke et al., 2021). Another study elaborates on the association of biomarkers such as t-tau with cognitive decline (Chen et al., 2021). The findings showed that higher levels of plasma biomarkers (i.e., Aβ42, t-tau, and Aβ42 × t-tau) were found in participants who showed a cognitive decline (the declined group) compared to those who did not (the stable group) and were associated with lower episodic verbal memory performance at baseline and a more significant annual decrease in MMSE score (Chen et al., 2021).

Instead of the classic post-viral syndrome, cognitive impairment may be a distinct post-COVID-19 manifestation caused by altered neuronal signaling in the brain due to the immune response triggered by the virus. However, there is no evidence of a link between inflammatory responses during acute infection (Woo et al., 2020). The presence of oligoclonal bands in one patient might be similar to the association of these bands in cognitive decline in other inflammatory and neurodegenerative diseases of the central nervous system like multiple sclerosis (Giedraitiene et al., 2021).

This study is limited by the number of studies available with the appropriate parameters. Most studies available were systematic and meta-analysis studies and observational studies that were inconclusive or unrelated to the topic of interest. Moreover, the majority of the excluded studies focused on neurological manifestations in general and did not necessarily mention cognitive dysfunction. Another limitation is the possibility that some relevant studies were not taken into account because they have been published in languages other than English (e.g., Chinese). We also did not have access to some other databases that may store some articles on COVID-19 and cognitive dysfunctions. And lastly, there could be some other studies on this theme in the literature that skipped our attention and analyses. However, a comprehensive search strategy that covers a broad range of evidence was implemented.

This systematic review gathered evidence that could provide clarity on the association between brain fog and COVID-19 infection. The information acquired in this study may help re-evaluate the impact of the virus. Furthermore, the use of the data gleaned from this analysis may assist in earlier treatment, allowing physicians and clinicians to manage the neurological manifestation effectively. Additionally, this will aid in the development of various therapeutic strategies to support COVID-19 patients in recovering from impaired cognitive capacity. Finally, the analysis of such data could provide an insight into the challenges that this virus could cause people in their prime years, particularly those in the workforce.

## CONCLUSION

Attention, memory, and executive function were the most frequently affected cognitive domains in COVID-19 patients. There was also a significant difference in the neuropsychological assessment scores between COVID-19 patients and HC. Interestingly, results from the included studies showed no correlation between cognitive dysfunction and depression. Increasing evidence suggests that cognitive dysfunction due to COVID-19 is manifested across disease severity ranging from asymptomatic to critical illness. The interplay of physical and cognitive impairments may lead to functional problems inhibiting health-related standards of life. The knowledge gained from this study may be used to improve the implementation of comprehensive treatment modalities and rehabilitation throughout the COVID-19 care continuum to remove such barriers and restore the meaningful lives of patients brought about by brain fog. The findings from this systematic review indicate multiple potential pathophysiological mechanisms related to cognitive dysfunction in COVID-positive individuals. Neuroinflammation is one of the mechanisms that have led to cognitive dysfunction based on the studies obtained. Neuroinflammation in the NFL protein and inflammatory levels indicated by CRP provide further insight into the pathophysiology that could lead to cognitive dysfunction. The evidence appears to be contrasting; however, from what was gathered, the CNS invasion is not the primary pathological mechanism due to the lack of studies that portray the presence of SARS-COV-2 concerning said mechanism. It is also suggested that neuroinflammation is not substantial enough even with the rising levels of pro-inflammatory markers due to the lack of value in numbers. Therefore, more studies are needed to substantiate these pathophysiological mechanisms further.

## Data Availability

Data are available on reasonable request. Extracted data are available on request to the corresponding author.

## Acknowledgment

The authors acknowledge the University of Santo Tomas College of Science, especially the Department of Biological Sciences, for the support and assistance.

## Authors’ Contributions

ADL conceptualized the study following discussion with JCD. MFDC, TRFM, OKSD, NDB, and AMOD designed the protocol, with feedback from ADL, ESB, ZGR, and JCD. MFDC, TRFM, OKSD, NDB, and AMOD ran the database search and oversaw the search, screening, full-text review and data extraction process. ZGR, ESB and ADL drafted the manuscript. All authors reviewed the draft, provided critical review and read and approved the final manuscript. The corresponding author, as guarantor, accepts full responsibility for the finished article has access to any data and controlled the decision to publish. The corresponding author attests that all listed authors meet the authorship criteria and that no others meeting the criteria have been omitted.

## Registration

This study is registered in PROSPERO (International Prospective Register for Systematic Reviews) as CRD42022325669.

## Author Note

If there is a need to amend the study protocol or results, the date of each amendment and the reason for the change will be described.

## Ethics statements

*Patient consent for publication*

Not applicable.

## Ethics approval

Ethical approval was not required for this systematic review, since all data came from information freely available in the public domain (i.e., published articles or conference abstracts). This study does not involve human participants.

