## Supplemental File No. 1 for "Clearing the Fog: A Systematic Review on Cognitive Dysfunction in COVID-19"

### Supplemental Appendix A. Search Strategy

**PubMed**
“COVID-19 and neurocognitive dysfunction” OR “brain fog”

**Oxford University Press**
“COVID-19 and neurocognitive impairment” OR “COVID-19 and brain fog” OR “COVID-19 and cognitive dysfunction” OR “COVID-19 and memory problems”

**ProQuest Health and Medical Complete**
“COVID-19 and brain fog” “COVID-19 and neurological mechanism”

**ScienceDirect**
“COVID-19 and neurocognitive impairment” OR “brain fog” OR “COVID-19 and confusion” OR “COVID-19 and memory problems” OR “COVID-19 and brain fog” OR “COVID-19 and pathophysiology” OR “COVID-19 and neurological mechanism” OR “COVID-19 and cognitive dysfunction”

**Jstor**
“COVID-19 and brain [public health]” OR “COVID and neurological impairment”

**HERDIN**
“COVID and brain fog”

**Google Scholar**
“Brain fog Covid-19” OR “brain fog covid-19” “Cognitive dysfunction Covid-19” OR “cognitive dysfunction covid-19” OR “Cognitive deficit Covid-19” OR “cognitive deficit covid-19” OR “neurocognitive dysfunction and Covid-19” OR “cognitive impairment Covid-19” OR “cognitive impairment COVID19”

**Cochrane Library**
“SARS-CoV-2 and neurocognitive impairment” OR “SARS-CoV-2 and brain fog” OR “COVID-19 and cognitive dysfunction”
